# IKONOS: An intelligent tool to support diagnosis of Covid-19 by texture analysis of x-ray images

**DOI:** 10.1101/2020.05.05.20092346

**Authors:** Juliana C. Gomes, Valter A. de F. Barbosa, Maira A. Santana, Jonathan Bandeira, Mêuser Jorge Silva Valença, Ricardo Emmanuel de Souza, Aras Masood Ismael, Wellington P. dos Santos

## Abstract

In late 2019, the SARS-Cov-2 spread worldwide. The virus has high rates of proliferation and causes severe respiratory symptoms, such as pneumonia. There is still no specific treatment and diagnosis for the disease. The standard diagnostic method for pneumonia is chest X-ray image. There are many advantages to using Covid-19 diagnostic X-rays: low cost, fast and widely available. We propose an intelligent system to support diagnosis by X-ray images. We tested Haralick and Zernike moments for feature extraction. Experiments with classic classifiers were done. Support vector machines stood out, reaching an average accuracy of 89.78%, average recall and sensitivity of 0.8979, and average precision and specificity of 0.8985 and 0.9963 respectively. The system is able to differentiate Covid-19 from viral and bacterial pneumonia, with low computational cost.

## 1 Introduction

A new kind of coronavirus cross the species barrier in December 2019 in China. This virus is related to severe acute respiratory syndrome coronavirus, for this reason it received the name SARS-Cov-2 [32, 36]. SARS-Cov-2 causes the coronavirus disease 19 (COVID-19) that affects the respiratory system causing several health complications as fever, cough, sore throat and in the most severe cases it can lead to shortness of breath and death.

Until the end of April 2020 the SARS-Cov-2 has spread from 213 countries infecting almost 3 million people worldwide and causing more than 200 thousand deaths [48]. Thus, there is a urgent need of methods to diagnosis the disease in a quick and precise way.

Due to the high virus spread rate it is necessary tests for diagnosis that are quick and precise [5]. The precise diagnosis in patients with Covid-19 permit they receive due medical attention, furthermore placing these patients in isolation will decrease the disease spread [5]. The test standard to the Covid-19 diagnosis is the quantitative polymerase chain reaction (qPCR), however this exam needs several hours to confirm positivity [20].

Rapid tests based on antibodies, such as IgM/IgG, are nonspecific for Covid-19, and may have very low sensitivity and specificity [9, 20, 21, 37, 47]. IgM/IgG detect the serological evidence of recent infection, not the presence of the virus. Besides it is not possible to ensure that the positive response is not from of antibodies of other coronaviruses and flu viruses [30]. According to Burog et al. [9] the use of IgM/IgG rapid test kits as definitive diagnosis of COVID-19 in currently symptomatic patients is not recommended. Liu et al. [33] concluded that IgM/IgG tests made in samples collected in the first week of illness have only 18.8% of sensitivity and 77.8% of sensibility. A low rate of sensitivity of IgM/IgG rapid test was found by Döhla et al. [20], they compare the results of IgM/IgG with reverse transcription polymerase chain reaction (RT-PCR) in 59 patients and conclude that the rapid test obtained 36.4% of sensitivity and 88.9% of specificity.

In the other hand IgG/IgM tests only reach high sensitivities and specificities when the viral charge is high. But it happens when the disease is in advanced levels [23, 26]. In the work of Liu et al. [33] they also concluded that tests performed during the second week of the disease have 100% of sensitivity and 50% of specificity.

Computed Tomography (CT) scans combine with RT-PCR has a great clinical value, considering that in CT images its possible to analyze the Covid-19 effects as bilateral pulmonary parenchymal ground-glass and consolidative pulmonary opacities [29]. However CT is a expensive exam that requires a dedicated room which can be an area of contamination among patients.

Thus, it is necessary the development of alternative forms of diagnosis. Recent works has been shown that chest X-ray image can be used to detect the Covid-19 with high accuracy [1, 2, 35, 45]. The uses of chest X-ray image in Covid-19 diagnosis is a good option seeing that it is the standard for diagnosing pneumonia and many establishments have it. Besides it is a quick exam and a has low price compared with others image exam like computed tomography and nuclear resonance magnetic.

In clinical practice, among the main pneumonia findings, it is common to find whitish or an opacity of one of the lungs when affected by pneumonia. These image artifacts are due to the production of mucus. Differently from pneumonias, Covid-19 is a disease that affects the blood, which can lead to thickening of the blood and thrombosis. Consequently, alveoli gas exchanges are impaired. Thus, before patients experienced breathing difficulties, it is common to notice changes in blood saturation. In this case, surfactant is damaged, leading alveoli to collapse and, therefore, compromising respiratory capacities. The tendency is both lungs be affected equally. Given the fact that the differential in the diagnosis is opacity, textures assume an important role in image diagnosis.

Machine Learning techniques has been using in several tasks including medical image classification [3, 4, 12, 13, 15–18, 31, 38–40, 42, 44, 46, 46]. Thus theses techniques can provides a secure and automatic way to diagnosis Covid-19 in chest X-ray images.

In this study, we propose an automatic system for Covid-19 diagnosis using machine learning techniques and chest X-ray images. In our experiments we used Multilayer Perceptron [25, 43], Support Vector Machine [7, 14], Random Trees, Random Forest [8, 22], Bayesian networks and Naive Bayes [10, 25]. As extractors attribute we used Haralick [24] and Zernike [27] extractors.

This work is organized as follows. Section 2 reviews related works in the diagnosis of Covid-19 using chest X-ray image. Section 3 presents dataset information and reviews the theoretical concepts necessary to understand the work. Section 4 shows the experimental results and the resulting desktop application we developped. 5 analyzes the experimental results. Section 6 ends the article with the conclusions.

## 2 Related Works

Some works have proposed the use of artificial intelligence for analysis of X-ray images and diagnosis of Covid-19. Narin et al. [35] developed a binary classifier to discriminate Covid-19 against and healthy patients). They tested three deep convolutional neural networks for classification: ResNet50, InceptionV3 and Inception-ResNetV2. In addition, they applied transfer learning techniques using ImageNet data. The reason for this choice is the small database. The models were made using 100 X-ray images, half of them is Covid-19 positive and the other half is healthy patients. The images were resized to 224 × 224 pixels. The experiments were performed using the Python language. For each configuration they tested 30 epochs, with 20% of data used for testing. Furthermore, they applied cross-validation with 5 folds. As performance metrics, the authors used accuracy, recall, specificity, precision and F1-Score. In this context, ResNet50 presented the best results. This model achieved an average accuracy of 98%, recall of 96% and precision of 100%.

Likewise, Apostolopoulos and Mpesiana [2] used Transfer Learning techniques with Convolutional Neural Networks for the diagnosis of Covid-19. However, Apostolopoulos and Mpesiana [2] developed multi-class classifiers. First, they used a base of 1427 X-ray images including cases of Covid-19, common bacterial pneumonia and healthy patients. Then, they included images of viral pneumonia for the development of a second classifier. All images were obtained through public repositories. The images were resized to 200 × 266 pixels. In some cases, to avoid distortion, they added a black background to achieve these dimensions. In this work, the CNNs tested were: VGG19, MobileNetV2, Inception, Xception and Inception ResNet v2. The training was conducted for ten epochs and tenfold cross-validarion. Considering the smaller database (without viral pneumonia), the best models found by the authors (VGG19 and MobileNet) have an average accuracy of 93.48% and 92.85%, respectively. For the most complete database, the average accuracy was 94.72%.

Apostolopoulos et al. [1] analyzed the problem of automatic classification of many lung diseases. The authors considered 7 classes: Covid-19, viral pneumonia, bacterial pneumonia, pleural effusion, chronic obstructive pulmonary disease and pulmonary fibrosis. The images were resized to 200 × 200 pixels and a black background added when necessary. Considering the small number of instances for each class, they applied data augmentation techniques. In this case, the images were randomly rotated or shifted horizontally or vertically. The work covered experiments with CNN Mobile Net v2, testing different learning strategies. 10-fold-cross-validation were used. Using the Of-the-Self-features learning strategy, the authors found an average accuracy of 51.98% and very low sensitivity and specificity. On the other hand, the best model was obtained with the Training-from-scratch strategy. The average accuracy found was 87.66%.

Sethy and Behera [45] also investigated radiographs as a diagnostic method for Covid-19. They organized two databases: the first, with 25 positive and 25 negative images for Covid-19 (or pneumonia); the second base, included MERS, SARS and ARDS in the Covid-19 positive class, with a total of 266 images. The authors examined the two bases using a deep learning method for extracting attributes (AlexNet, VGG16, VGG19, GoogleNet, ResNet18, ResNet50, ResNet101, InceptionV3, InceptionResNetV2, DenseNet201 and XceptionNet) and SVM as a classifier. As in other works, transfer learning techniques were applied. The experiments were carried out in Matlab, using the deep learning toolbox. In addition, they performed 100 independent simulations for each configuration. The work showed that ResNet50 with SVM presented the best results with an average accuracy of 95.38%.

As for the diagnosis of Covid-19, each of the proposals mentioned above has advantages and disadvantages. They are summarized in the following table. In the last line, it can observed the characteristics of this present work.

**Table 1:** Summary of the works described in this section

| Work | Propose | Advantages | Disadvantages |
| --- | --- | --- | --- |
| Narin et al. (2020) | Binary classification (Covid-19 positive and healthy patient) using Deep CNN and Transfer learning methods | High performance with mean accuracy of 98% | Small database and the work does not include other respiratory diseases |
| Apostolopoulos and Mpesiana (2020) | Multi-classes classification (Covid-19, healthy, bacterial and viral pneumonia) using CNN and Transfer Learning methods | Mean accuracy of 94%, even with the inclusion of viral and bacterial pneumonia | Authors did not comment on the computational cost of the solution, which is usually high when using CNNs |
| Apostolopoulos et al. (2020) | Multi-classes classification with 7 classes (Covid-19, viral and bacterial pneumonia, pulmonary edema, pleural effusion, chronic obstructive pulmonary disease and pulmonary fibrosis) | The model includes many respiratory diseases and it achieves 87.66% mean accuracy | Authors did not comment on the computational cost of the solution, which is usually high when using CNNs |
| Sethy and Behera (2020) | Binary classification using deep learning and SVM | They reached an accuracy of 95.38% in Covid-19 diagnosis | Small database. In addition, authors included SARS, MERS, and ARDS cases in Covid-19 positive class, which does not help in medical decision |
| **This work** | AI system with multi-classes classification (Covid-19, healthy, viral and bacterial pneumonia) | The system can diagnosis Covid-19 with 89.78% of mean accuracy. Its prototype is already developed and it has low computational cost | System does not include a large amount of respiratory diseases |

## 3 Methods

### 3.1 Proposed method

In this context, this work proposes the development of IKONOS, a desktop application to support and optimize the diagnosis of Covid-19 through chest X-ray images. We also aim to produce a tool for easy maintenance and scalability, using algorithms of low computational complexity. Thus, we seek to provide one more diagnostic method to combat the current pandemic, in order to complement this process and minimize costs.

The basic functioning of this system is this: the medical team of the health institution must request chest X-ray examinations from patients with symptoms characteristic of Covid-19 After receiving the digital images, the radiologist or healthcare professional can then upload the image to the application. The images will then be analyzed by an intelligent system. It will be able to carry out multi-classes classification, differentiating multiple respiratory diseases such as Covid-19, viral pneumonia and bacterial pneumonia. For this, machine learning techniques will be used aiming at good results, even if training the system with a small set of real images. The methods tested in this work will be: Haralick and Zernike for feature extraction and multiple classical classifiers will be tested and compared. Finally, the system will provide a diagnosis, which can be viewed on the computer screen. The diagnosis will be available with accuracy, sensitivity and specificity information, so that the health professional can make the decision of the subsequent clinical conduct. This proposal is summarized in Figure 1.

**Fig. 1:**
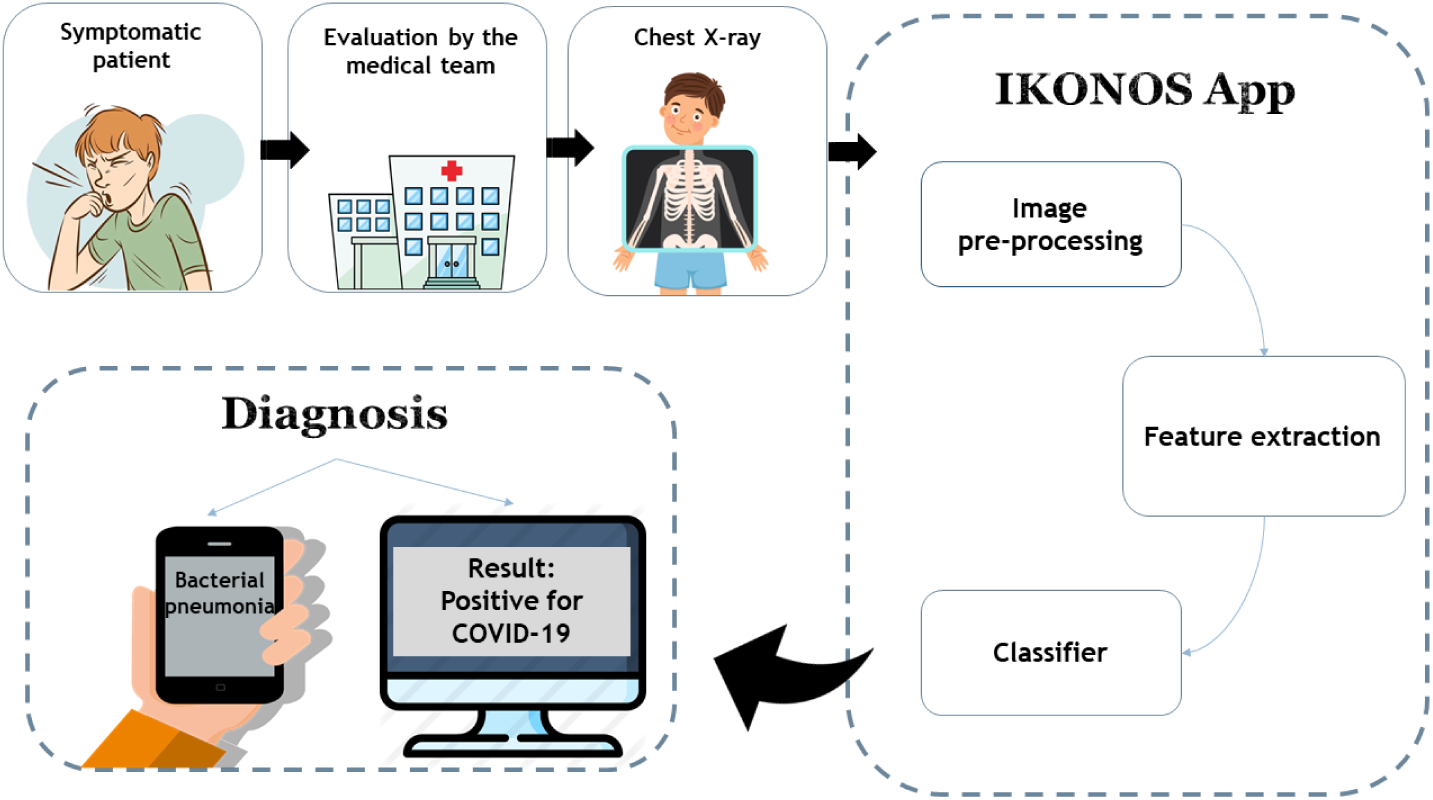
Diagram of the proposed method. Chest X-rays of symptomatic patients can be loaded into the IKONOS app. The application consists of an intelligent system capable of extracting features and classifying the image among 4 cases: healthy patients, viral pneumonia, bacterial pneumonia and Covid-19. The result can be viewed on any computer with software installed. Accuracy, sensitivity and specificity information will also be available, helping the professional in decision making.

### 3.2 Dataset

For the development of this project, we used X-ray images from different databases. Images of viral pneumonia, bacterial pneumonia and healthy patients were obtained on the Kaggle website, available in competition format by Paul Mooney [34]. On the other hand, radiographs of patients with Covid-19 were obtained from 5 different databases:

– Open source GitHub repository shared by Dr. Joseph Cohen [11]. This group built the database with images from publications as they are images that are already available.
– Covid-19 database, made available online by Societa Italiana di Radiologia Medica e Interventistica [19]
– Radiopaedia database
– Database from Peshmerga Hospital Erbil

All the patients classify with Covid-19 in our database were diagnosed with gene sequencing and reverse transcription polymerase chain reaction (RT-PCR). The joining of the bases resulted in 6350 images. The number of images for each of the classes is described in the Table 2 below. Figure 2 presents sample images of healthy patients, viral pneumonira, bacterial pneumonia, and Covid-19.

**Table 2:** Number of X-ray images of each class: Covid-19, viral pneumonia, bacterial penumonia, and healthy patients

| Class | Total |
| --- | --- |
| Covid-19 | 464 |
| Viral Pneumonia | 1490 |
| Bacterial Pneumonia | 2783 |
| Healthy patients | 1583 |

**Fig. 2:**
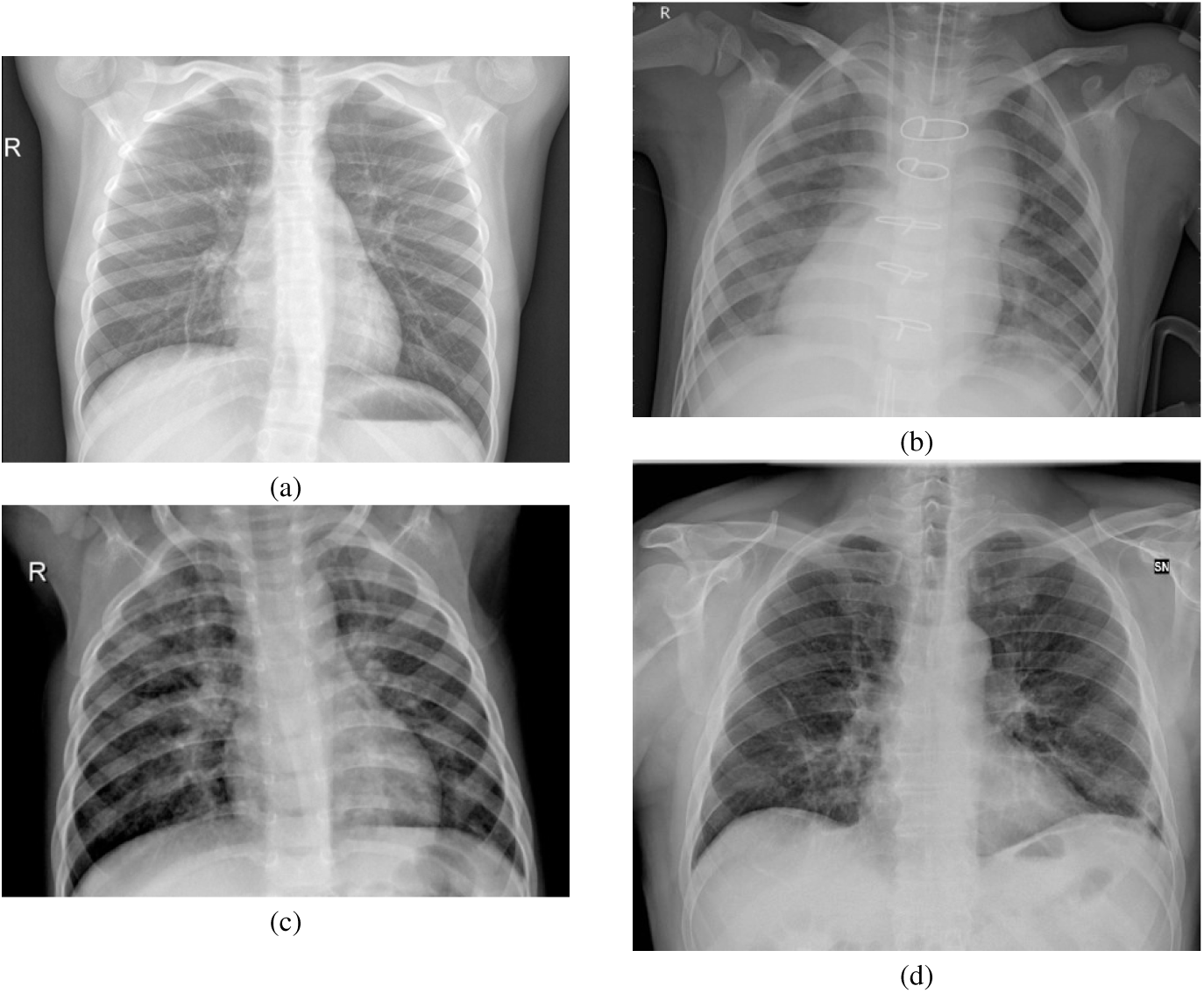
Sample images of the datasets used in this work: healthy lungs (a), bacterial pneumonia (b), viral pneumonia (c), and Covid-19 (d).

There are some limitations to the databases that we used for this project. First, Covid-19 images have no information about the severity of the patients. Thus, we believe that it is possible that the images are of more severe cases.

In addition, the images of bacterial and viral pneumonia are relatively old, and not from this year of the pandemic (2020). This means that patients diagnosed with pneumonia did not have symptoms characteristic of Covid-19. We also emphasize that we do not have patient demographic information, such as sex, age and presence of cormobilities.

### 3.3 Feature extraction: Haralick and Zernike

The descriptor of Haralick extracts feature related to the textures of the images. Texture is an intrinsic property of surfaces. It contains important information about their structural composition. From Haralick’s moments, it is possible to differentiate textures that do not follow a certain pattern of repetition throughout the image. This method calculate statistical information associated to the co-occurrence matrices from the gray scale image. These matrices show the occurrence of certain pixel intensities. Each *p*(*i*, *j*) of the matrix consists on the probability of going from one pixel of intensity *i* to another pixel of intensity *j*, according to a certain distance and an angle of the neighborhood [24].

In this way, each matrix considers the relationship between a reference pixel and its neighborhood. Thus representing the spatial distribution and dependence of gray levels in regions of the image. In this study, we considered two versions of the image to perform the feature extraction. The first was the gray scale image and the second was a pre-processed image, using Kohonen maps as filter. This process resulted in 104 features per image.

The Zernike descriptor is another widely used tool for feature extraction. It extract information related to shape or geometry from an image. Zernike moments are invariant to rotation, not redundant and robust to noise [27]. To calculate these moments, we consider the center of the image as the center of a unit disk. The moments are calculated from the projections of the intensity function of an image on the orthogonal base functions. So we calculate each of the 64 moments from the Zernike family of polynomials, *V_n,m_*, described by Equations 1 and 2.

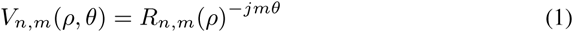

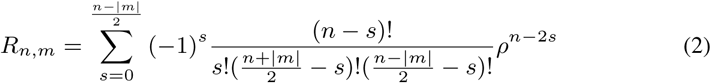

The 64 descriptors are equally divided into two groups, according to the polynomial order (*n*). To calculate the features, *n* and *m* from Equations 1 and 2 assume the values in Table 3.So,at the end of the process, we have 32 moments of low order and 32 of high order. Shape-related features of an image are also relevant in the context of identifying pathologies, since these conditions usually result in changes in geometric patterns.

**Table 3:** Moments of Zernike according to group and parameters *n* and *m*

| Group | n | m | Number of moments |
| --- | --- | --- | --- |
| 1 | 3 | 1,3 | 32 |
|  | 4 | 0,2,4 |  |
|  | 5 | 1,3,5 |  |
|  | 6 | 0,2,4,6 |  |
|  | 7 | 1,3,5,7 |  |
|  | 8 | 0,2,4,6,8 |  |
|  | 9 | 1,3,5,7,9 |  |
|  | 10 | 0,2,4,6,8,10 |  |
| 2 | 10 | 2,6,10 | 32 |
|  | 11 | 3,7,11 |  |
|  | 12 | 0,4,8,12 |  |
|  | 13 | 1,5,9,13 |  |
|  | 14 | 2,6,10,14 |  |
|  | 15 | 3,7,11,15 |  |
|  | 16 | 0,4,8,12,16 |  |
|  | 17 | 1,5,9,13,17 |  |

### 3.4 Classification

In this section we briefly discussed the machine learning methods used to classify X-ray images.

#### 3.4.1 Multilayer Perceptron

The psychologist Rosenblatt [43] was one of the pioneers in the concept of artificial neural networks. In 1958 he proposed the perceptron model for supervised learning. Perceptron is the simplest form of neural network used for binary classifications of linearly separable patterns. It consists of a single neuron with adjustable synaptic weights and a bias [25].

Multilayer Perceptron (MLP) is a generalization of the single layer perceptron. It consists of a feed-foward network with an input layer, hidden layers and one output layer. The addition of hidden layers allows to the network the ability to classify more complex problems than single layer perceptron such as image classification [4, 28, 41].

The main algorithm for training MLPs is the error backpropagation algorithm. Based on a gradient gradient, backpropagation proceeds in two phases: propagation and back propagation. In the propagation phase, an output is obtained for a given input pattern. In back progation phase, an error is calculate using the desired output and the output obtained in the previous phase. Then the error is used to update the connection weights. Thus backpropagation aims to iteratively minimize the error between the network output obtained and the desired output [25].

#### 3.4.2 Support Vector Machine

Created by Vapnik [7, 14] the support vector machine (SVM) perform a nonlinear mapping on the dataset in a space of high dimension called feature space. So a linear decision surface, called hyperplane, is constructed in order to separate distinct classes [14].

Thus, the training process of a support vector machine aims to find the hyperplane equation which maximizes the distance between it and the nearest data point. That hyperplane is called optimal hyperplane [25].

#### 3.4.3 Decision Trees

Decision trees are a type of supervised machine learning model. They are widely used to solve both classification and regression problems. In general, trees have nodes, which are structures that store information. In a tree there are basically four types of nodes: root, leaf, parent and child. The root node is the starting point and has the highest hierarchical level. One node may connect to another, establishing a parent-child relationship, in which a parent node generate a child node. Leaf nodes, in turn, are terminal nodes, so they have no children, and represent a decision. In this way, using such trees, the algorithm makes a decision after following a path that starts from the root node and reaches a leaf node. There are several types of decision trees. They usually differ from the way the method goes through the tree structure. Among these types, the methods Random Tree and Random Forest are two of the main ones.

Random Tree algorithm uses a tree built by a stochastic process. This method considers only a few features in each node of the tree, which are randomly selected [22]. The Random Forest algorithm, in turn, consists in a collection of trees. These trees hierarchically divide the data, so that, each tree votes for a class of the problem. At the end, the algorithm choose the most voted class as the prediction of the classifier [8].

#### 3.4.4 Bayesian Network and Naive Bayes

Bayes Net and Naive Bayes are classifiers based on Bayes’ Decision Theory. Bayesian classifiers, also called the test procedure by the Bayes hypothesis, seek to find a minimum mean risk. By considering a set of correct decisions and a set of incorrect decisions, they use conditional probability to create the data model. The product of the frequency of each decision and the cost involved in making the decision are the weights [25]. For a Gaussian distribution, Bayesian networks behave like a linear classifier. Its behavior is comparable to that of a single layer perceptron.

In the standard Bayes Network algorithm, it assesses the probability of occurrence of a class from the values given by the others. So, this method assumes dependence between the features. Naive Bayes, on the other hand, considers that all features are independent of each other, being only connected to the class [10]. This method does not allow dependency between features. Since this assumption represents an unrealistic condition, the algorithm is considered "naive".

#### 3.4.5 Parameters settings of the classifiers

All experiments were performed using the Weka software. For each configuration described in the table below 25 simulations were performed, using 10-fold cross-validation. The parameters used in each machine learning method is shown in Table 4.

**Table 4:** Classifiers parameters: SVMs with linear, 2-degree and 3-degree polynomials, and RBF kernels; MLPs with 50 and 100 neurons in the hidden layer; random forests with 10 to 100 trees; random tree; and standard Byesian networks and Naive Bayes classifiers.

| Classifier | Parameters |
| --- | --- |
| SVM | Kernel Linear<br>Kernel Polynomial E2<br>Kernel Polynomial E3<br>Kernel RBF |
| MLP | 50 neurons in the hidden layer<br>100 neurons in the hidden layer |
| Random Forest | Number of trees: 10, 20, 30, 40, 50, 60,<br>70, 80, 90, 100<br>Batch size: 100 |
| Bayesian networks | Batch size: 100 |
| Naive Bayes | Batch size: 100 |
| Random tree | Batch size: 100<br>Seed = 1 |

### 3.5 Metrics

In order to analyze the classification performance, we used six metrics: Accuracy, Recall, Sensitivity, Precision, Specificity, and Kappa Index. Accuracy assesses the proportion of images correctly classified on all results. It can be calculated according to Equation 3. In a machine learning context, the term Recall is common. However, in the medical world, the use of the sensitivity metric is more frequent.

Mathematically, both terms are the same. They are the rate of true positives, and indicate the system’s ability to correctly detect people who are sick (with Covid-19, for example). They can be calculated according to Equation 4. Precision, on the other hand, is the fraction of the positive predictions that are actually positive. The precision can be calculated according to Equation 5.

Specificity is the metric that evaluates a model’s ability to predict true negatives of each available category or the rate of true negatives. This means that specificity indicates the classifier’s ability to correctly exclude healthy or disease-free people. It can be calculated as the Equation 6. Finally, the Kappa index is a very good measure that can handle very well both multi-class and imbalanced class problems, as the one proposed here. It can be calculated according to the Equation 7. These four metrics allow to discriminate between the target condition and health, in addition to quantifying the diagnostic exactitude [6]. These metrics are described as following:

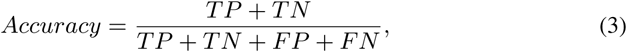

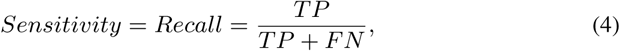

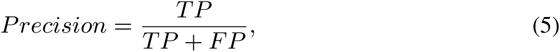

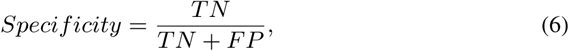

where TP is the true positives, TN is the true negatives, FP is the false positives, and FN the false negatives.

The *κ* coefficient (kappa) is defined as follows:

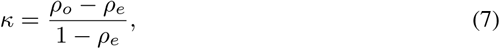

where *ρ_o_* is observed agreement, or accuracy, and *p_e_* is the expected agreement, defined as following:

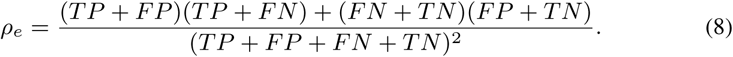

## 4 Results

After feature extraction, two databases were generated: the first using Haralick; and the second, using Haralick and Zernike. Both bases were trained using multiple classic machine learning methods.

### 4.1 Classifiers experiments results

#### 4.1.1 Results using Haralick for feature extraction

Figure 3 shows boxplots referring to the four metrics for evaluating the classifiers. Considering the database with Haralick moments, the SVM polynomial with exponent 3 presented the best result in all metrics. In contrast, networks based on Bayes’ theory showed the worst results. Table 5 shows the comparison of these two cases.

**Fig. 3:**
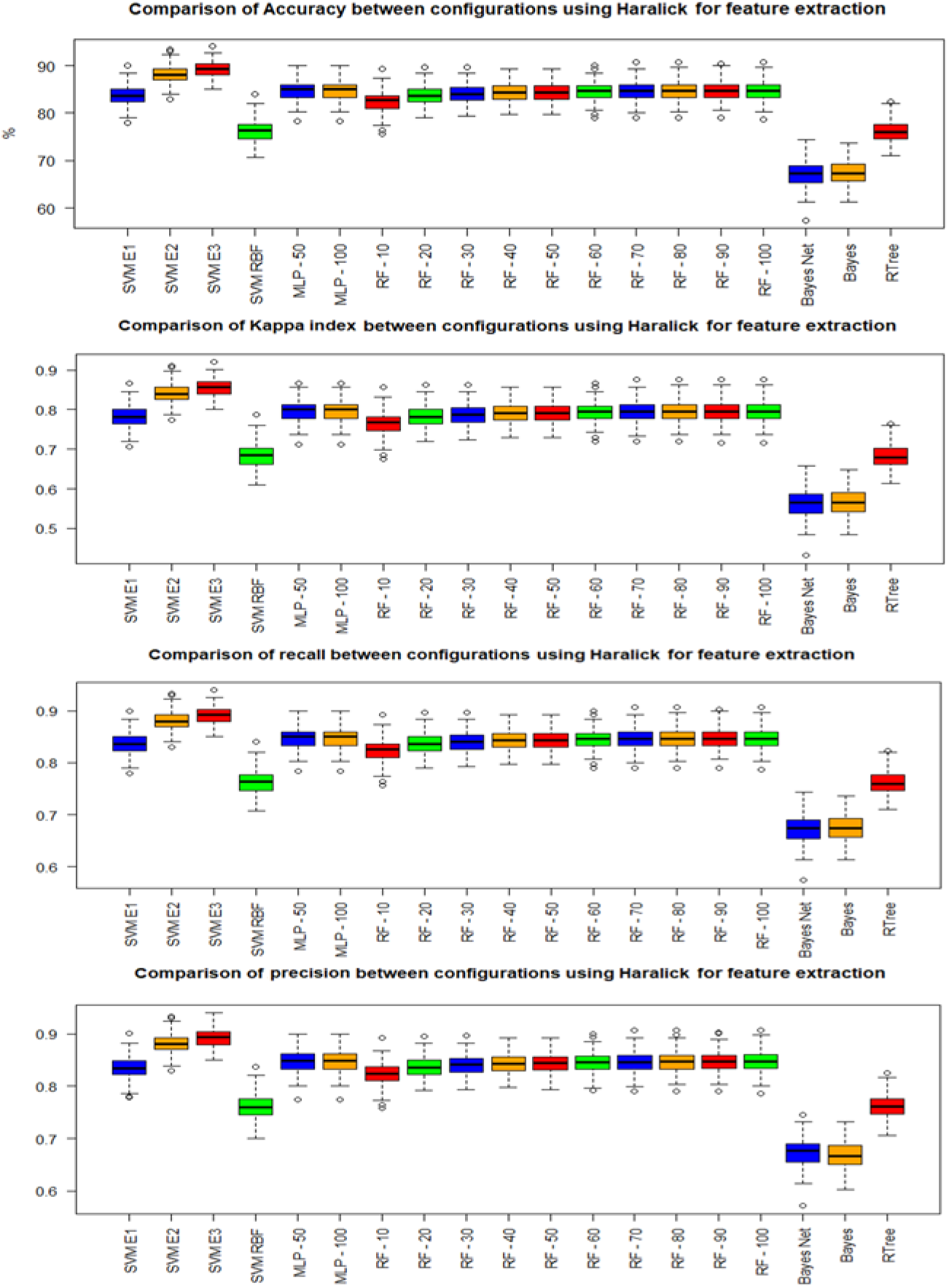
Boxplots with results for all tested classifier

**Table 5:** Summary of the results using Haralick.

| Best and worst models using Haralick for feature extraction |  |  |  |  |  |  |  |  |  |  |  |  |
| --- | --- | --- | --- | --- | --- | --- | --- | --- | --- | --- | --- | --- |
| Model | Accuracy |  | Kappa Index |  | Recall |  | Precision |  | Sensitivity |  | Specificity |  |
|  | Mean | StDev | Mean | StDev | Mean | StDev | Mean | StDev | Mean | StDev | Mean | StDev |
| SVM poly - E3 | 89.1387 | 1.7105 | 0.8552 | 0.0228 | 0.8914 | 0.017 | 0.8924 | 0.017 | 0.8914 | 0.017 | 0.9934 | 0.0054 |
| Bayesian net | 67.1933 | 2.4952 | 0.5626 | 0.3327 | 0.6719 | 0.025 | 0.6786 | 0.0253 | 0.6719 | 0.025 | 0.9453 | 0.0157 |

Table 6 shows precision, recall, specificity, and sensitivity results for the best model. Notice that Covid-19 is well-discriminated, presenting precision of 0.979, recall of 0.977, specificity of 0.993, and sensitivity of 0.977. Table 7 presents the confusion matrix. Bacterial and viral pneumonias cause more confusion. 143 images of viral pneumonia were classified as bacterial pneumonia, and 125 images of bacterial pneumonia were classified as viral pneumonia.

**Table 6:** Results obtained with Haralick moments: precision, recall, specificity, and recall results for each class, i.e. Covid-19, bacterial pneumonia, viral pneumonia, and healthy lungs. Notice that Covid-19 is well-discriminated, presenting precision of 0.979, recall of 0.977, specificity of 0.993, and sensitivity of 0.977.

| Class | Precision | Recall | Specificity | Sensitivity |
| --- | --- | --- | --- | --- |
| Covid-19 | 0.979 | 0.977 | 0.993 | 0.977 |
| Bacterial pneumonia | 0.796 | 0.817 | 0.93 | 0.817 |
| Viral pneumonia | 0.813 | 0.795 | 0.939 | 0.795 |
| Healthy | 0.981 | 0.979 | 0.994 | 0.979 |

**Table 7:** Results obtained with Haralick moments: Confusion matrix showing that bacterial and viral pneumonias cause more confusion. 143 images of viral pneumonia were classified as bacterial pneumonia, and 125 images of bacterial pneumonia were classified as viral pneumonia. The Covid-19 classification, on the other hand, has a high hit rate.

| a | b | c | d | classified as |
| --- | --- | --- | --- | --- |
| 733 | 6 | 7 | 4 | a → Covid-19 |
| 9 | 613 | 125 | 3 | b → Bacterial pneumonia |
| 4 | 143 | 596 | 7 | c → Viral pneumonia |
| 3 | 8 | 5 | 734 | d → Healthy |

#### 4.1.2 Results using Haralick and Zernike for feature extraction

Figure 4 on the other hand, shows boxplots with the results obtained with the second database, using moments from Haralick and Zernike. As in the previous case, the Support vector machines showed the best results. However, in this case, the best performance was using degree 2. Furthermore, the Bayesian network presented again the worst performance. Table 8 summarizes these results.

**Fig. 4:**
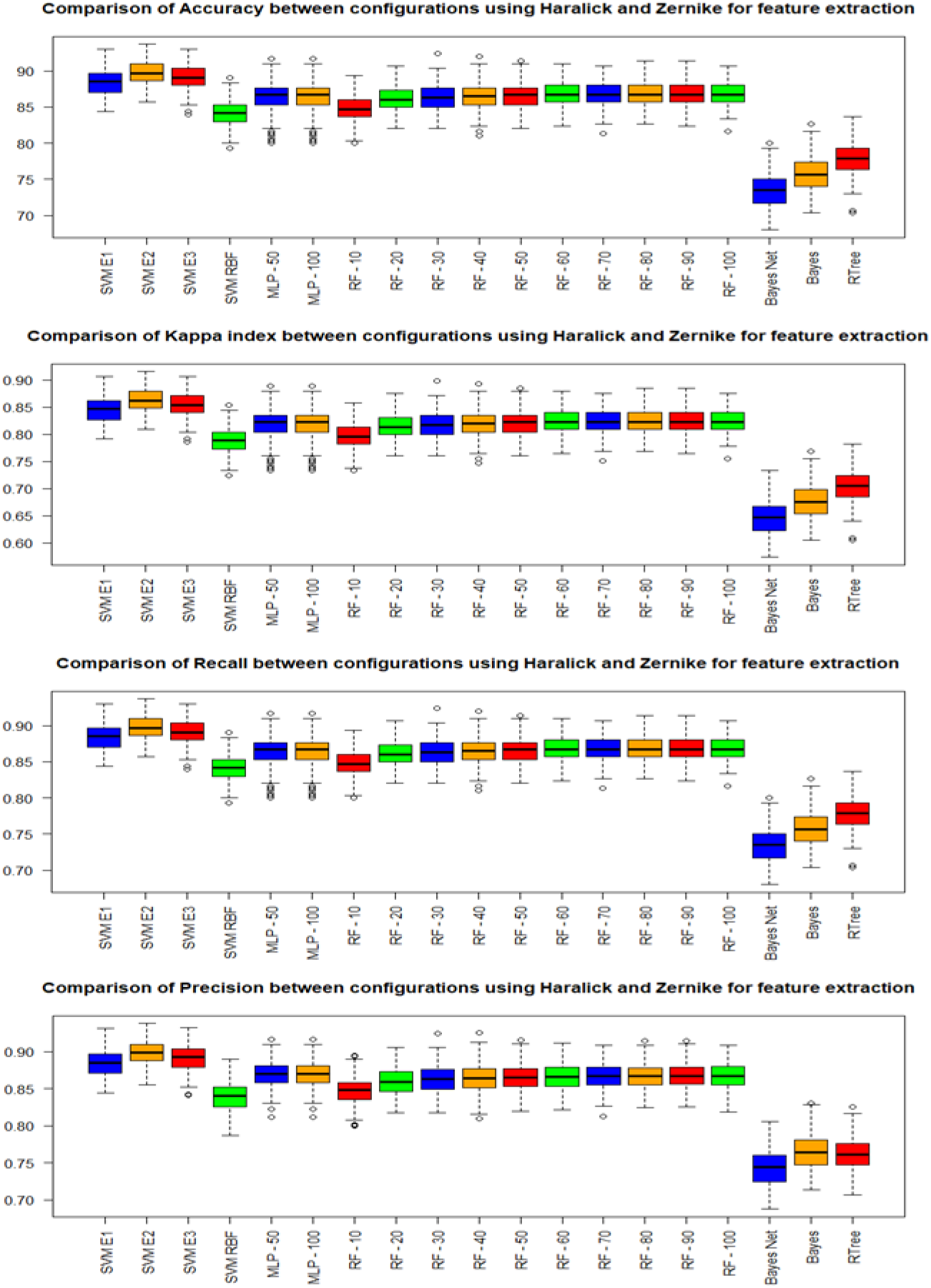
Boxplots with results for all tested classifier

**Table 8:**
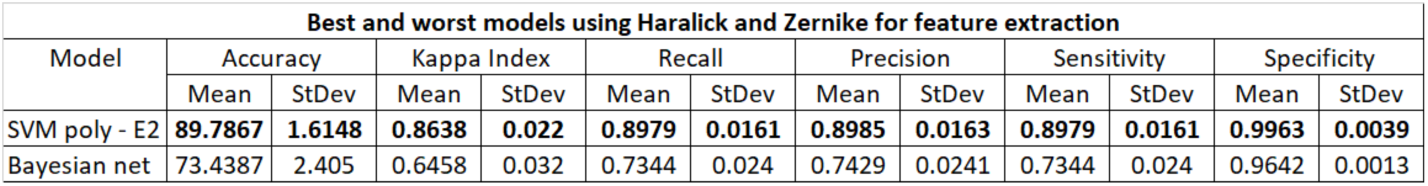
Summary of the results using Haralick and Zernike

| Best and worst models using Haralick and Zernike for feature extraction |  |  |  |  |  |  |  |  |  |  |  |  |
| --- | --- | --- | --- | --- | --- | --- | --- | --- | --- | --- | --- | --- |
| Model | Accuracy |  | Kappa Index |  | Recall |  | Precision |  | Sensitivity |  | Specificity |  |
|  | Mean | StDev | Mean | StDev | Mean | StDev | Mean | StDev | Mean | StDev | Mean | StDev |
| SVM poly - E2 | 89.7867 | 1.6148 | 0.8638 | 0.022 | 0.8979 | 0.0161 | 0.8985 | 0.0163 | 0.8979 | 0.0161 | 0.9963 | 0.0039 |
| Bayesian net | 73.4387 | 2.405 | 0.6458 | 0.032 | 0.7344 | 0.024 | 0.7429 | 0.0241 | 0.7344 | 0.024 | 0.9642 | 0.0013 |

Table 9 presents results obtained with Haralick and Zernike extractors: precision, recall, specificity, and sensibility results for each class, i.e. Covid-19, bacterial pneumonia, viral pneumonia, and healthy. It shows recall and sensitivity of 0.991, precision of 0.988 and specificity of 0.996 for Covid-19. Table 10 presents the confusion matrix for Haralick and Zernike moments, showing that bacterial and viral pneumonias cause more confusion in the classification.

**Table 9:** Results obtained with Haralick and Zernike extractors: precision, recall, specificity, and sensibility results for each class, i.e. Covid-19, bacterial pneumonia, viral pneumonia, and healthy. It shows recall and sensitivity of 0.991, precision of 0.988 and specificity of 0. 996 for Covid-19. These results highlight the considerable ability of the proposed approach to discriminate Covid-19 between the other classes.

| Class | Precision | Recall | Specificity | Sensitivity |
| --- | --- | --- | --- | --- |
| Covid-19 | 0.988 | 0.991 | 0.996 | 0.991 |
| Bacterial pneumonia | 0.806 | 0.853 | 0.932 | 0.853 |
| Viral pneumonia | 0.822 | 0.777 | 0.944 | 0.777 |
| Healthy | 0.969 | 0.963 | 0.99 | 0.963 |

**Table 10:**
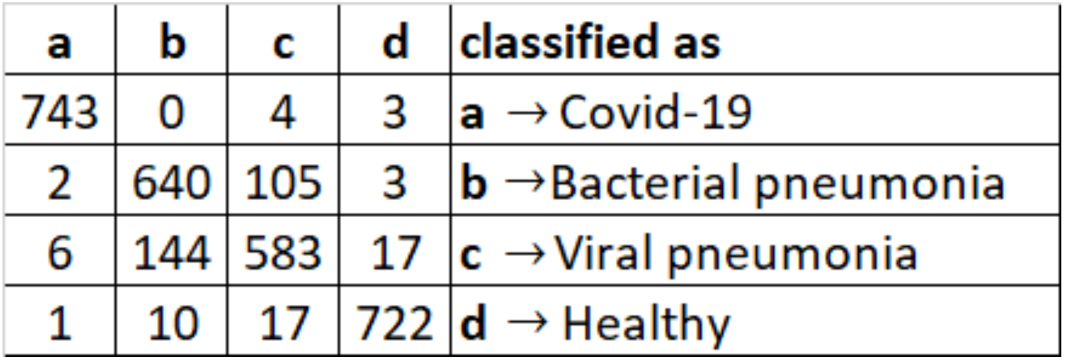
Confusion matrix for Haralick and Zernike moments, showing that bacterial and viral pneumonias cause more confusion in the classification. 143 images of viral pneumonia were classified as bacterial pneumonia, and 105 images of bacterial pneumonia were classified as viral pneumonia. Covid-19 classification, on the other hand, reached a considerably high hit rate.

| a | b | c | d | classified as |
| --- | --- | --- | --- | --- |
| 743 | 0 | 4 | 3 | **a** → Covid-19 |
| 2 | 640 | 105 | 3 | **b** → Bacterial pneumonia |
| 6 | 144 | 583 | 17 | **c** → Viral pneumonia |
| 1 | 10 | 17 | 722 | **d** → Healthy |

### 4.2 Desktop interface

The prototype of the developed system is fully functional in a desktop version. It works like this: digital X-ray images can be loaded into the software from any computer, as shown in Figure 5 below. They are analyzed by the intelligent system, capable of performing multi-class classification.

**Fig. 5:**
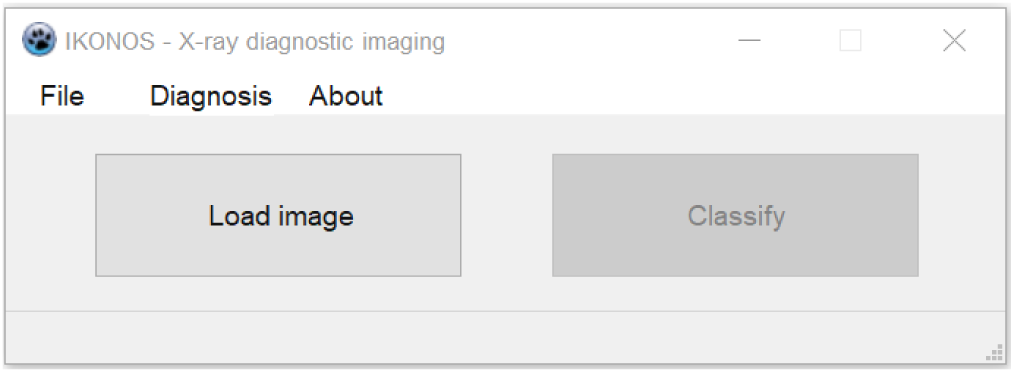
IKONOS App interface

The result can be visualized by the medical team through the computer, as shown in Figure 6. Accuracy, precision and recall information are also available. These allow them to confirm their initial hypotheses, or else to outline others, for the continuation of this investigative process. Considering the demand for a real-time solution, the intelligent model has low associated computational cost, that is, low memory consumption and adequate execution time.

**Fig. 6:**
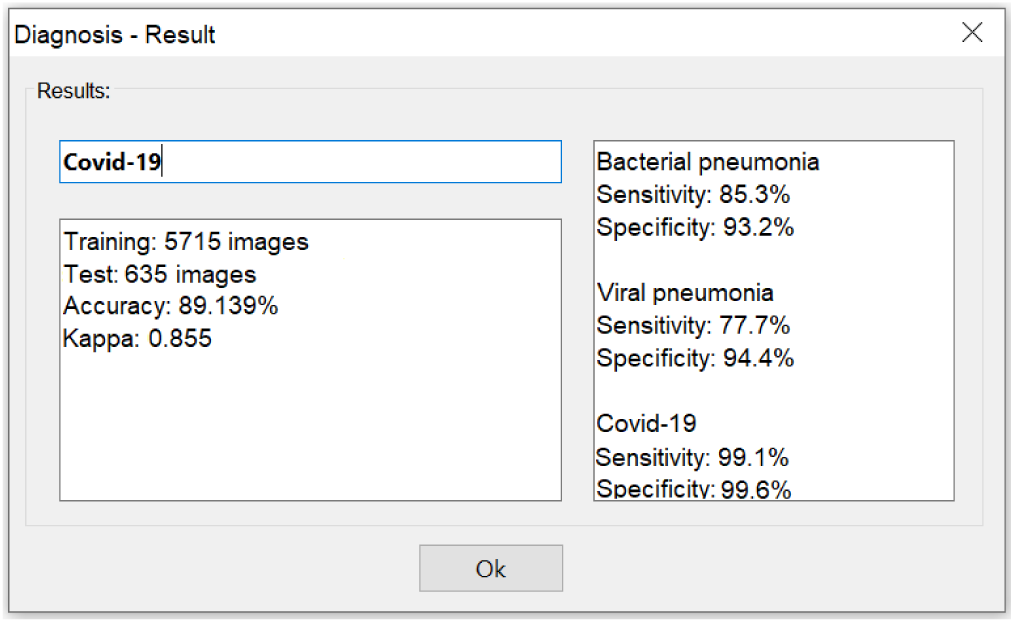
IKONOS App - Results screen

The code for this desktop version is freely available for non-commercial purpose on Github repository: github.com/Biomedical-Computing-UFPE/Ikonos-X-Desktop.

## 5 Discussion

When analyzing the box plots and mean and standard deviation values, as can be seen on Figure 3 and Table 5, we see that the SVM-E3 presented both higher mean and significantly lower standard deviation values. These results point to the possibility of classifying the four selected classes using classical methods and with low computational cost.

Taking into account precision, recall, specificity, and sensitivity results for the best model, as seen on Table 6, we can perceive that Covid-19 is well-discriminated, presenting precision of 0.979, recall of 0.977, specificity of 0.993, and sensitivity of 0.977. Looking at the confusion matrix on Table 7, it is clear that bacterial and viral pneumonias cause more confusion. 143 images of viral pneumonia were classified as bacterial pneumonia, and 125 images of bacterial pneumonia were classified as viral pneumonia. It highlights the difficult to discriminate the two major kinds of pneumonia. The Covid-19 classification, on the other hand, reached a higher hit rate. Comparing the second approach, i.e. the combination of Haralick and Zernike moments on the box plots of Figure 4, we can see the Support vector machines showed the best results as well. However, in this case, the best performance was using 2-degree polynomial kernels. Furthermore, the Bayesian network presented again the worst performance, as can be seen on Table 8.

For the combination of Haralick and Zernike moments, we obtained recall and sensitivity of 0.991, precision of 0.988 and specificity of 0.996 for Covid-19, as can be seen on Table 9. These results highlight the considerable ability of the proposed approach to discriminate Covid-19 between the other classes. The confusion matrix on Table 10 shows that bacterial and viral pneumonias cause more confusion in the classification. 143 images of viral pneumonia were classified as bacterial pneumonia, and 105 images of bacterial pneumonia were classified as viral pneumonia. The Covid-19 classification, on the other hand, reached a considerably high hit rate.

When we compared the results obtained with SVM for Haralick and for moments of Haralick and Zernike, we noticed a small improvement in the metrics in the second case. There is both an increase in averages and a decrease in standard deviations. Despite this, the difference between the two cases is not considerable. This situation indicates that the simplest method, using feature extraction with Haralick and SVM, may be sufficient for a good classification, with high sensitivity and specificity for Covid-19 image diagnosis.

## 6 Conclusion

Our motivation with this work is to save lives, through a correct diagnosis of Covid-19. We know that X-rays are one of the steps in the Covid-19 diagnostic process in most of the affected countries, and also the standard exam for assessing pneumonia. In addition, with the support of artificial intelligence, this technique shows results of recall, precision, specificity and sensitivity much higher than those of rapid tests.

Another motivation of this work was to evaluate texture and shape descriptors. In clinical practice, it is common to find whitish or an opacity of one of the lungs when affected by pneumonia. This is due to the production of mucus. Differently, Covid-19 is a disease that affects the blood, which can lead to thickening of the blood and thrombosis. As a consequence, the gas exchanges carried out at the level of the alveoli in the lungs are impaired. Therefore, before patients experienced breathing difficulties, it is common to notice changes in saturation. In this case, surfactant is damaged, leading alveoli to collapse and, therefore, reducing respiratory capacities. The tendency is both lungs be affected equally, unlike traditional pneumonias. Thereby, knowing that the differential in the diagnosis is opacity, we are talking about textures, emphasizing the importance of the studied descriptors.

In this way, using these descriptors for feature extraction with classic classifiers, we were able to develop an intelligent system with low computational cost. This factor is fundamental when we work with the perspective of a system accessed by several health units simultaneously. Thus, this work achieved relevant results with an evident computational cost much lower than those using deep learning techniques.

These results lead us to believe that clinical diagnosis aligned with image diagnosis could be sufficient for the diagnosis of Covid-19 and to proceed to treatment, although samples should be sent for molecular diagnosis in parallel for epidemic purposes. Evidences indicate lung changes could be found in X-ray images in the very beginning of Covic-19 symptoms. The combination of X-ray image diagnosis and clinical diagnosis can accelerate the beginning of treatment. Consequently, thanks to the wide availability and ease of X-ray equipment, many lives can be saved.

## Data Availability

All results were based on publicly available Covid-19 x-ray images datasets.

## Acknowledgements

The authors are grateful to the Federal University of Pernambuco and the Brazilian research agencies FACEPE, CAPES and CNPq, for the partial financial support of this research.

## Compliance with Ethical Standards

This study was funded by the Federal University of Pernambuco and the Brazilian research agencies FACEPE, CAPES and CNPq.

All procedures performed in studies involving human participants were in accordance with the ethical standards of the institutional and/or national research committee and with the 1964 Helsinki declaration and its later amendments or comparable ethical standards.

